# Measuring Institutional Research Culture: An Analysis of Accepted Abstracts at the Academic Surgical Congress

**DOI:** 10.1101/2024.05.21.24307727

**Authors:** Tejas S. Sathe, Ojas S. Sathe

## Abstract

**Introduction:** Understanding which institutions have a strong research culture is important for trainees, but existing metrics such as NIH funding do not always reflect a trainee’s lived experience. Here, we present a descriptive analysis of accepted abstracts at the Academic Surgical Congress (ASC).

**Methods:** We analyzed accepted abstracts at the Academic Surgical Congress between 2016 and 2024. We identified institutions with the most accepted abstracts, overall and by year. We also identified institutions with the greatest year-over-year improvement between 2022 and 2024. Finally, we calculated associations between accepted abstracts and NIH funding and also between accepted abstracts and leadership in the ASC’s hosting societies.

**Results:** A total of 12,142 abstracts were accepted by the ASC between 2016 and 2024. The University of Alabama and the University of Michigan led overall abstract acceptances and also led in most years. Baylor and Massachusetts General Hospital had the greatest absolute and percent year-over-year increase in abstract acceptances in the post-COVID era. There was a positive association between NIH funding and abstract acceptances. Among the top 40 institutions by accepted abstracts in 2024, institutions in the top 10 were over 10 times as likely to have leadership representation in the societies that host the ASC.

**Conclusions:** Conference abstracts may serve as a useful measure of trainee-relevant research culture. Trainees can utilize this information when selecting residency programs, and programs can use this information to optimize trainee research experiences.

## Introduction

What institutions are “good” at surgical research? Whatever “good at research” means exactly, it seems to be a metric that prospective applicants gauge when choosing residency programs and institutions optimize for when recruiting and promoting faculty. Nevertheless, it is unclear how to measure this metric in a manner that is practically relevant to trainees. Historical measures of academic success such as research funding from the National Institutes of Health (NIH) may be less relevant in an era when trainees are increasingly pursuing scholarly work in disciplines such as surgical education and surgical innovation, possibly under the guidance of mentors with non-federal research funding or even no funding. For example, though one researcher may dramatically impact an institution’s research profile through a large NIH grant, this effect may only be practically relevant to the few trainees that work in that researcher’s laboratory. Perhaps a more impactful question for prospective surgery applicants is rather whether an institution promotes a culture of research that permeates the lived experience of trainees.

We hypothesize that accepted conference abstracts may serve as a potential measure of trainee-relevant research productivity, and thus, institutional research culture. Academic conferences are an important venue for sharing novel research, but also have other benefits, such as creating opportunities for networking and offering pathways to publish peer-reviewed manuscripts. Furthermore, simply presenting at a conference implies other administrative corollaries to a strong research culture. For example, if an institution has multiple trainees presenting abstracts, then one can infer that such an institution has the requisite financial resources to support registration and travel expenses and can maintain appropriate clinical coverage to accommodate brief absences from work. To study this topic further, we aim to analyze abstracts accepted to the Academic Surgical Congress (ASC), a national, annual surgical meeting co-hosted by the Association for Academic Surgery (AAS) and the Society of University Surgeons (SUS). Few prior studies analyzed ASC abstracts, usually to study author demographics, calculate how many of the abstracts were published, or measure the impact of those resulting publications. However, these studies often involved the manual review of small subsets of presented abstracts - such as those accepted for plenary presentations.^1–3^ To date, no study has performed a comprehensive analysis of institutional representation across all accepted ASC abstracts.

In this paper, we present an analysis of accepted ASC abstracts. Specifically, we present descriptive data on which institutions achieved the most accepted abstracts and also which institutions were able to increase their number of accepted abstracts in a short time period. Next, we study the association between accepted abstracts and institutional NIH funding and also the association between accepted abstracts and the number of institutional faculty that serve in leadership roles in the societies that host the ASC. Through this study, we aim to propose conference abstracts as a useful measure of an institution’s research culture. Moreover, we hope that our data can inform efforts to expand research opportunities for trainees and that our analytic approaches can be useful to future researchers.

## Methods

### Data Sources

We obtained data on accepted abstracts from the Academic Surgical Congress ranging from 2016 to 2024. The dataset contained information on the session type (e.g. QuickShot, Oral, or Plenary), abstract type (e.g. Basic Science, Clinical/Outcomes, or Education), scientific area (e.g. Health Informatics, COVID-19, Immunology/Transplantation), and clinical area (e.g. Endocrine, Cross Disciplinary). In addition to the title and body text, each abstract contained an author block containing all of the authors and their institutional affiliations.

We separately obtained data on National Institutes of Health (NIH) funding received by academic surgery programs from the Blue Ridge Institute for Medical Research, a non-profit institution that provides data on NIH funding to individual investigators and institutions.^4^ Data was obtained for 2016-2023.

### Obtaining Institution Data

For each abstract, we split the author block into two sections comprising the authors and institutions. If an abstract was associated with more than one institution, we extracted the first institution in the list to correspond with the primary affiliation of the first author. While exploring the dataset we found that there was no standardized methodology for writing the institutional affiliation. For example, one abstract contained the affiliation “University Of Alabama at Birmingham, Birmingham, Alabama, USA” while another contained the affiliation “University Of Alabama at Birmingham, Department Of Surgery, Birmingham, Alabama, USA”. Intuiting that these two affiliations were actually the same, we used the following techniques to standardize the data.

First, we split each institutional affiliation into a list of comma-delimited phrases (e.g. “University of Alabama at Birmingham”, “Department of Surgery”, “Birmingham”, “Alabama”, and “USA”). From this list, we extracted the first element that contained the phrase “university”, “hospital”, “institute”, or “medical center”. If none of these phrases occurred in the institution list, we extracted the first element.

Next, we found that there were several institutional affiliations that were similar without being an exact match. For example, our extraction resulted in institutional affiliations like “University of Alabama” and “University of Alabama Birmingham”, and “University of Alabama at Birmingham”, likely referring to the same institution. To address this, we first removed filled words like “or” or “at” as well as the initial keywords “university”, “hospital”, “institute”, or “medical center”. We then performed fuzzy string matching using a threshold of 90% similarity to resolve these differences. We manually reviewed that data for the top 20 institutions in terms of overall accepted abstracts and resolved errors as needed. Of note, we had to manually combine some institutions that are the same but can be described using non-similar words. One example of this is “Hospital of the University of Pennsylvania” and “Perelman School of Medicine”, which represent the same institution. Following these three rounds of review, we arrived at an institutional affiliation corresponding to the first author of each abstract. A schematic of our approach is shown in Figure 1.

**Figure 1:**
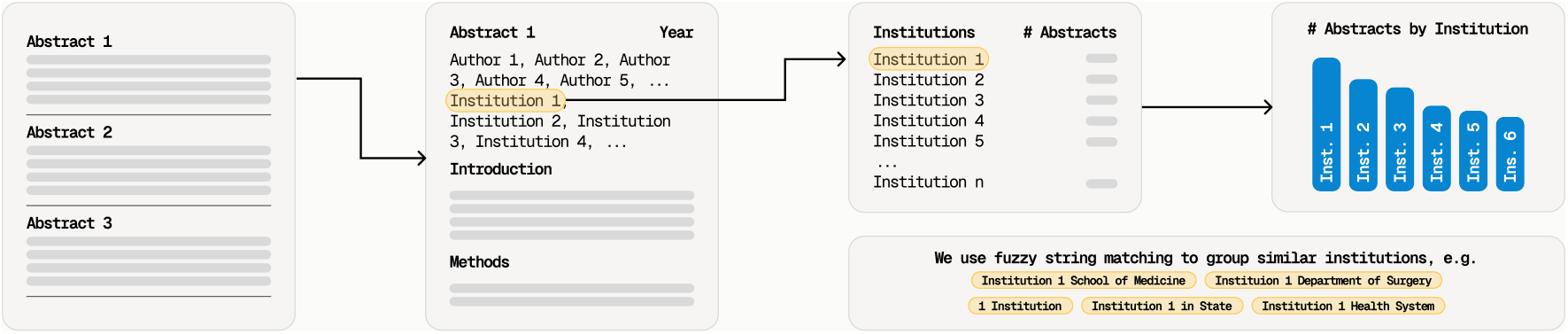
Schematic of our approach to identifying the institutional affiliation of abstract first authors using fuzzy string matching. Using this technique, we estimated the number of abstracts per institution.

### Descriptive Abstract Data

We included abstracts with no missing data in the author block and excluded duplicate abstracts. First, we calculated the total number of accepted abstracts and the number of accepted abstracts by year. We also broke this data down by session type and abstract type. We then calculated the total number of accepted abstracts by institution, both overall and by year. For the latter, we calculated abstracts both as an absolute number and as a percentage of total conference abstracts. For example, if an institution had 50 abstracts and a total of 1000 abstracts were accepted, that institution represented five percent of the conference’s total abstracts. We felt that this calculation could be used as a marker of institutional prominence in a given year. We plotted the above metrics for the top 20 institutions by overall abstracts between 2016 and 2024.

### Year-Over-Year (YoY) Changes in Accepted Abstracts

Next, we wanted to understand how the number of abstracts changed between consecutive years. For this analysis, we used a subset of our initial dataset, including only the top 20 institutions by overall abstract acceptance and the years 2022 to 2024 to avoid capturing any changes that may have resulted acutely from the COVID-19 pandemic. We then measured both the absolute and percent difference in accepted abstracts between 2023 and 2022 and between 2024 and 2023 for each of the top 20 institutions. We plotted the top ten positive absolute and percent changes in YoY abstract acceptance in bar plots.

### Association Between NIH Funding and Accepted Abstracts

As NIH funding data for 2024 was not available, 2024 abstract data was excluded from this analysis. In order to examine whether NIH funding was associated with accepted ASC abstracts, we performed several analyses. First, we measured the Spearman’s rank correlation coefficient between NIH funding and accepted abstracts for each year between 2016 and 2023. Then, we performed a linear regression analysis with log-transformed average of NIH funding as the independent variable and accepted abstracts as the dependent variable. We chose to log-transform NIH funding due to the wide variance in NIH funding and to avoid biasing the regression against institutions with significant funding. From this, we obtained an R2 score, F-statistic, and p-value.

### Association Between Leadership Representation and Accepted Abstracts

We queried the websites of the AAS and SUS to identify surgeons who held leadership positions within these societies. ^5,6^ For the AAS, we included all surgeons who had a role other than ‘member’ or ‘member candidate’. This included leading officers such as the president, president-elect, and secretary as well as committee co-chairs. For the SUS, we included the executive council and representatives. We performed a Google Search to obtain the institutional affiliation of each AAS leader and obtained SUS leadership institutional affiliations directly from the website.

For our analysis, we utilized abstracts from 2024 to coincide with the timeframe when our identified society officers were in their active terms. We further only included the top 40 institutions by 2024 abstract count. We considered that an institution had leadership representation if one or more surgeons from that institution held a leadership position within the AAS or SUS. We categorized the top 10 institutions by 2024 abstract count (within the top 40 cohort) as top performers. To measure the association between leadership representation and abstract performance, we calculated an odds ratio.

We then wanted to understand whether there was a dose-response relationship between the number of active officers within the AAS and SUS and abstract count, so we performed a linear regression of these two variables within this subset of top 40 institutions by 2024 abstract count.

### Data and Code

All analyses were completed in Python 3.10. We used ChatGPT Plus with Code Interpreter to perform a preliminary analysis of the data and suggest analytical techniques. We used GitHub Copilot within Microsoft VS Code to write our actual code. The code and dataset are available on GitHub.

## Results

### Descriptive Abstract Data

A total of 12,142 abstracts were accepted by the Academic Surgical Congress in the years 2016-2024. Accepted abstracts increased from 2016 (n=1,125) to 2020 (n=1,742), then fell to a nadir in 2022 (n=848). After this, they rose to a high point in 2024 (n=1,746). Of note, the COVID-19 pandemic emerged immediately after the 2020 conference and the 2021 conference was held virtually. The ASC returned as an in-person conference in 2022 and onward (Figure 2).

**Figure 2:**
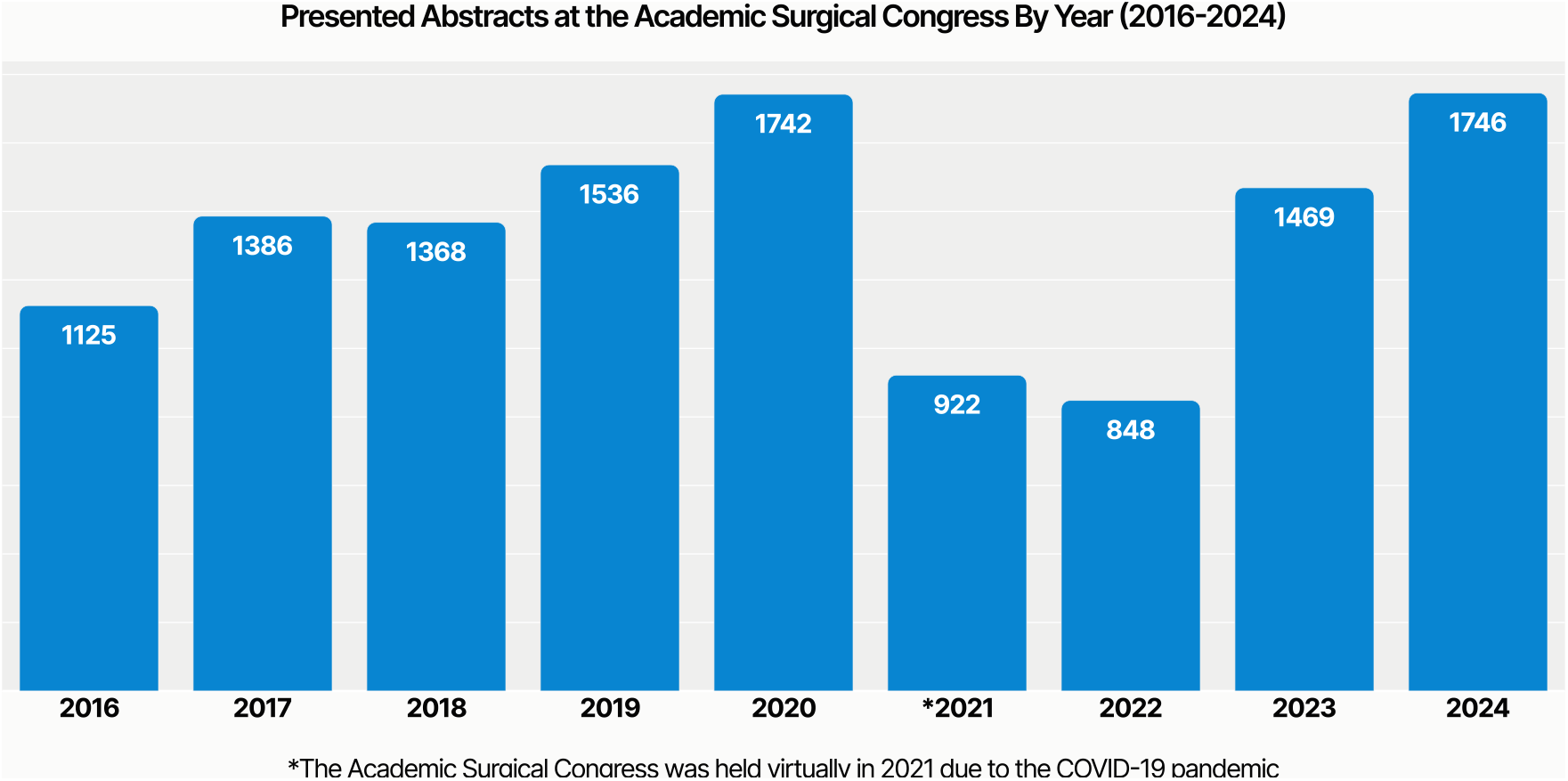
Number of overall accepted ASC abstracts between 2016 and 2024.

### Abstracts by Institution and Year

The University of Alabama led in overall accepted abstracts (n=589), followed by the University of Michigan (n=429). The remainder of the top 20 institutions had between 170 and 256 abstracts (Figure 3). When grouping the data by year, the results were similar. While The University of Michigan led in accepted abstracts in 2016 (n=42), the University of Alabama led in all other years, reaching a high point in 2020 (n=94). The University of Michigan had the second most abstracts in all of the years that the University of Alabama held the lead except 2020, when it placed third (n=47) behind Johns Hopkins (n=59) (Figure 4A). When analyzing accepted abstracts by institution as a percentage of total accepted abstracts each year, the relative relationships between institutions remained the same. In this analysis, the University of Alabama and the University of Michigan both achieved their peak percentages in 2022 (8% vs 5%, respectively) (Figure 4B).

**Figure 3:**
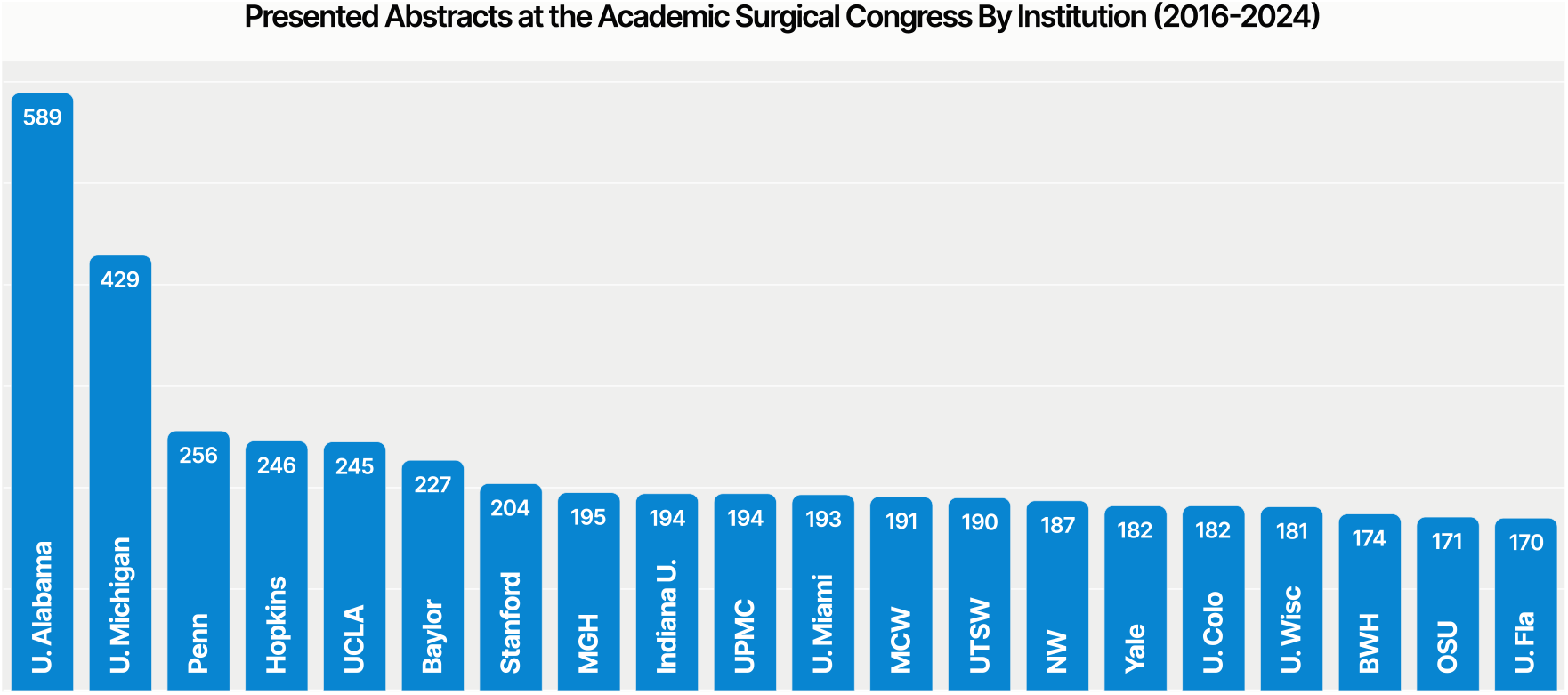
Number of accepted ASC abstracts by institution between 2016 and 2024. U. Alabama = The University of Alabama at Birmingham, U. Michigan = The University of Michigan, Penn = Hospital of the University of Pennsylvania, Hopkins = Johns Hopkins University, UCLA = The University of California Angeles, Baylor = Baylor College of Medicine, Stanford = Stanford University, MGH = Massachusetts General Hospital, Indiana U. = Indiana University, UPMC = University of Pittsburgh Medical Center, U. Miami = The University of Miami, MCW = Medical College of Wisconsin, UTSW = University of Texas Southwestern, NW = Northwestern University, Yale = Yale University, U. Colo = The University of Colorado, U. Wisc = The University of Wisconsin-Madison, BWH = Brigham and Women’s Hospital, OSU = The Ohio State University, U. Fla = The University of Florida.

**Figure 4:**
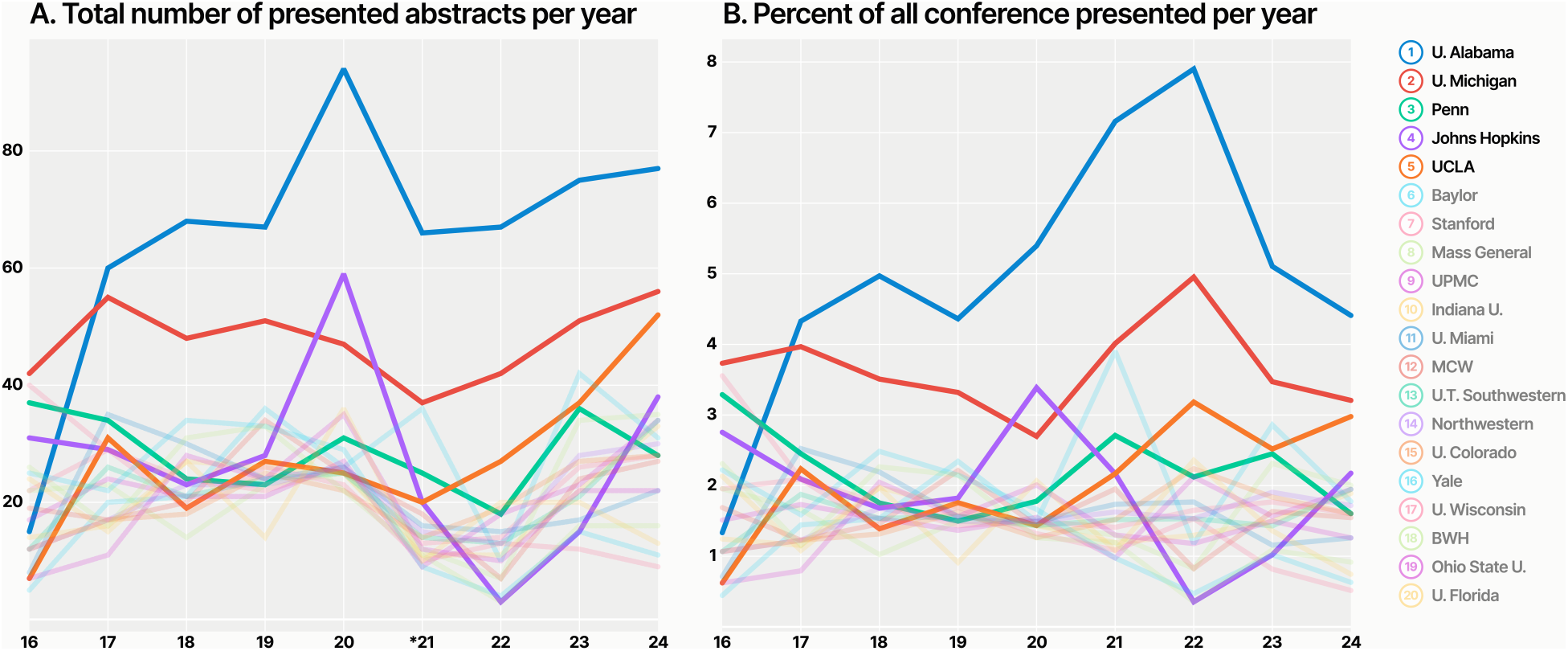
Number of accepted ASC abstracts by institution by year between 2016 and 2024 in absolute terms (A) and as a percent of overall abstracts for that year (B). The opacity of institutions 6-20 has been reduced for improved clarity.

### Year-Over-Year (YoY) Changes in Accepted Abstracts

Between 2022-2024, Baylor had the largest YoY increase in accepted abstracts in 2023 (Δ=32), followed by Massachusetts General Hospital in 2023 (Δ=27), and Johns Hopkins in 2024 (Δ=23) (Figure 5A). Brigham and Women’s Hospital had the greatest YoY percent increase in accepted abstracts in 2023 (Δ=433%), followed by Johns Hopkins in 2023 (Δ=400%), and Massachusetts General Hospital in 2023 (Δ=386%) (Figure 5B). Further data on YoY changes is shown in Figure 5.

**Figure 5:**
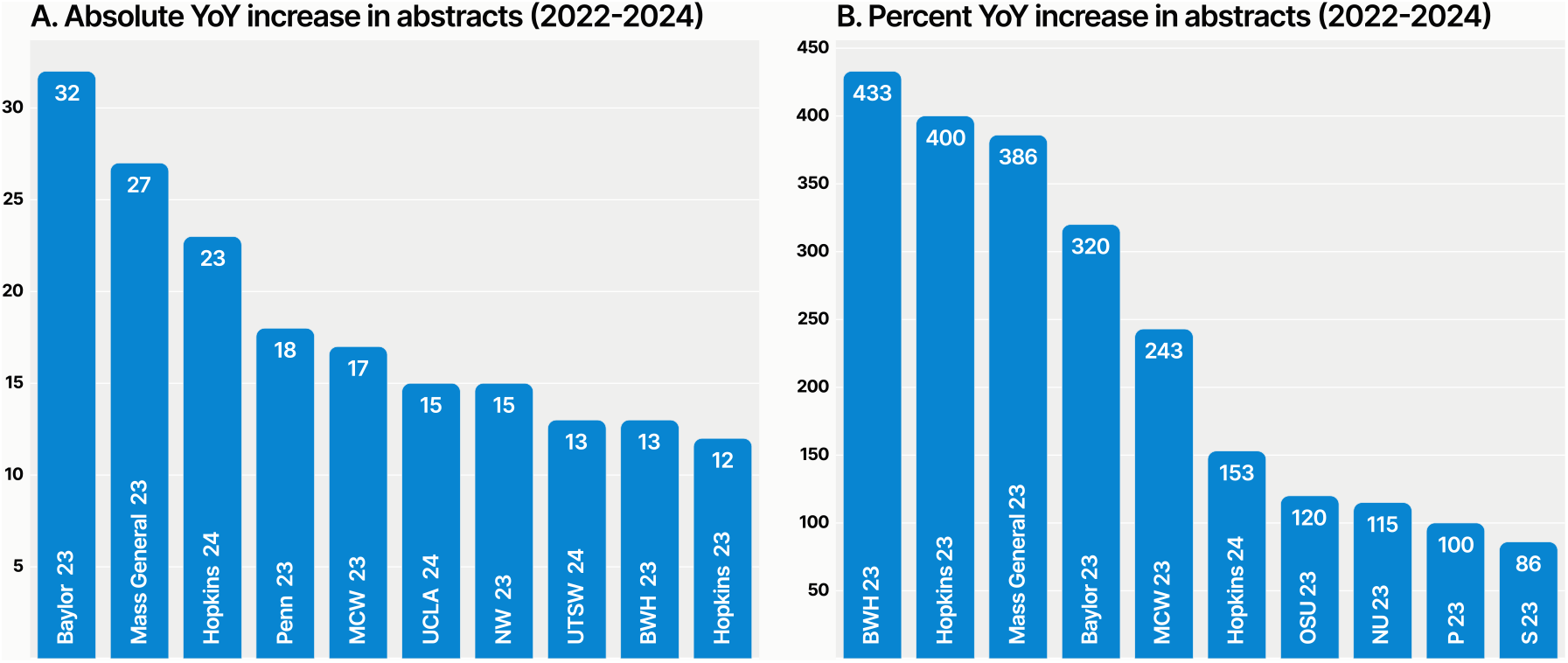
Absolute (A) and Percent (B) year-over-year changes in ASC accepted abstracts between 2022 and 2024, reflecting the post-COVID era.

### Association between NIH Funding and Accepted Abstracts

We obtained data on NIH Funding for surgery programs in each year between 2016 and 2023. For institutions that received both NIH funding and had accepted abstracts at the ASC, the Spearman’s rank correlation coefficient was between 0.450 and 0.550 for each year, and all correlations were statistically significant with a p-value of less than 0.001 (Supplementary Table 1).

When performing a linear regression between the log-transform of average NIH funding and total accepted abstracts, the model demonstrated that log-NIH funding accounted for approximately 31% of the variance in the number of accepted abstracts (R^2^ = 0.307), with an F-statistic of 41.6 (p *<* 0.001). However, our results were complicated by statistically significant Omnibus and Jarque-Bera tests, both of which suggested a non-normal distribution of residuals (actual minus predicted accepted abstracts), which is a key assumption of linear regression. We corroborated the positive association between log-NIH funding and abstracts using other regression models, but present here the linear regression for simplicity. We visualized this data with a scatter plot showing the overlaid regression line and highlighting both overperformers in blue and underperformers in red, respectively (Figure 6).

**Figure 6:**
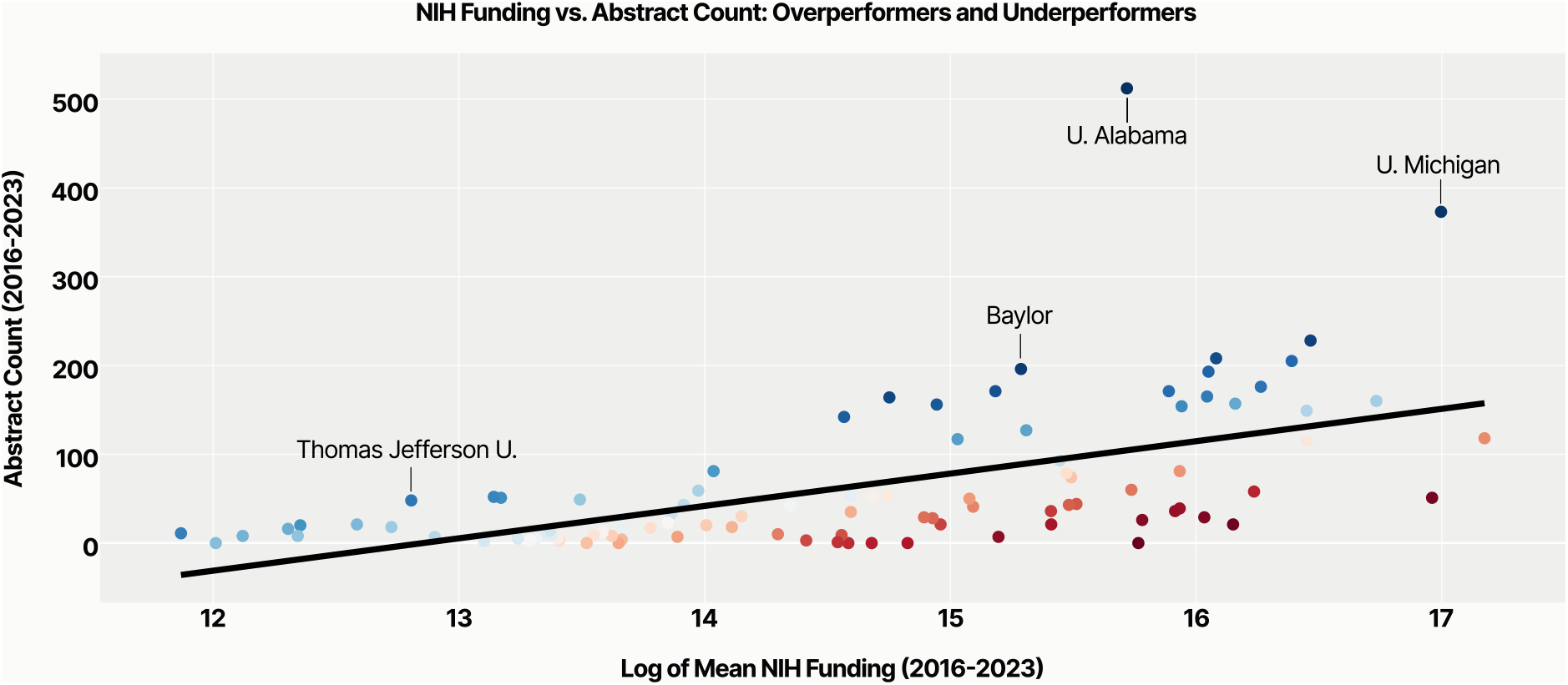
Linear regression between the log-transform of mean NIH funding and total accepted ASC abstracts between 2016 and 2023. Data points are colorized based on the percentile of the residual between the actual and predicted number of abstracts. Overperformers are shown in blue while underperformers are shown in red. Darker colors indicate greater absolute values of residuals.

### Association between Leadership Positions and Accepted Abstracts

In our subset of top 40 institutions by 2024 abstract count, we calculated an odds ratio of 11.8 (p=0.013) between being in the top 10 in accepted abstracts and having leadership representation. In other words, institutions in the top quartile of this already selective cohort were over ten times as likely to have officers in either the AAS or SUS leadership than institutions in the next 30. We found that Spearman’s rank correlation coefficient between number of abstracts and number of leadership positions was 0.50 (p=0.001) for institutions in the top 40 accepted abstract count in 2024 (Supplementary Table 2).

## Discussion

In this study, we found that the number of accepted abstracts at the Academic Surgical Congress is increasing. We also found that two institutions, the University of Alabama at Birmingham and the University of Michigan, led abstract acceptances overall and also in each year we studied. On the other hand, institutions such as Baylor, Massachusetts General Hospital, Johns Hopkins, and Brigham and Women’s Hospital showed large increases in abstract acceptance between consecutive years, showing that these institutions were able to improve their conference participation dramatically in a short period of time.

We found that an institution’s NIH funding was positively associated with the number of accepted abstracts. Of note, both correlations were moderately positive and significant, suggesting that this positive association is real but not insurmountable. Of note, many of the top 20 institutions (such as the University of Alabama and the University of Michigan) were also overperformers, which means that more abstracts are accepted from these institutions than would be predicted by our model based on their NIH funding. But there were also other institutions outside of the top 20, like Thomas Jefferson University and Georgetown University that overperformed their NIH funding. Of note, as the regression coefficient was only 0.31, almost 70% of variability in abstract count is explained by other, and possibly more easily modifiable factors.

We also found that institutions at the very top of abstract productivity are over ten times as likely to have representation within the AAS or SUS leadership. This dramatic odds ratio raises interesting additional questions. Do institutions have both leadership representation and strong abstract performance because of some external confounder? After all, both are measures of academic success. Or does having a leadership position in one of these societies lead surgeons to encourage their trainees to submit their research to the ASC? Or alternatively are residents who had positive experiences at the ASC working at the same institutions at which they trained and achieving leadership positions within the ASC’s organizing societies? Perhaps, some combination of all of the above is in effect.

What is ultimately most interesting to us is how this data can be used to help programs improve their academic productivity. For example, if you are a faculty at a program that does not have strong ASC representation and wants to improve it, what is the best thing you can do? Win an NIH grant? Chair an AAS committee? A useful next step of our work would be to interview stakeholders at top performing institutions, those that improve from year to year, or those that overperform quantitative metrics such as NIH funding. From talking to these individuals, we may be able to develop a playbook that all institutions, students, and trainees can benefit from in the future.

There are several limitations to this study. The largest is our inability to determine any causal relationships from the associations presented. Moreover, it is possible that our fuzzy string matching was not perfect. We attempted to address this by doing a manual review of institutions and making corrections as needed. We did this exhaustively for the most commonly represented institutions, but it was not feasible to do so for the whole dataset. It is thus possible that our numbers are not exact, but any errors should be small enough as to not alter the overall message of our data. An additional limitation was our choice to only consider the institutional affiliation of the first author. This may have resulted in underestimation in participation levels for institutions that were involved in multi-institutional studies. Another source of error inherent to our study design was the ability to sample data from only one national conference. While the Academic Surgical Congress is a large and popular conference for trainees, it is only one of many conferences at which trainees can present research. As a result, an analysis of the ASC alone does not represent a complete picture of trainee academic productivity, and it is quite possible that our institutional rankings would differ if we had data on other conferences.

Here, we demonstrate the first comprehensive institutional analysis of accepted abstracts at a large, national surgical conference. This data can be useful for many stakeholders within academic surgery. Medical students can use it to identify institutions that are academically productive in a trainee-relevant manner. We argue that while NIH funding may provide a sense of an institution’s overall research prowess, conference acceptances are more relevant to a trainee’s lived experience. Second, residents and faculty surgeons can use this data to inform quality improvement projects. By comparing one’s institutional data to others, institutions can set metrics to improve research productivity. Finally, data like this can be useful to conference organizers, who can draw from it to understand who is presenting at their meetings. Our hope is that this data may be used to optimize transparency around institutional research cultures and promote a more positive research environment for trainees.

## Data Availability

All data produced are available online at: https://github.com/tsathe/asc-abstract-proj/

## Acknowledgments

We thank Jason Levine from BSC Management, Inc. for providing abstract data.

## Supplementary Tables

**Supplementary Table 1:** Spearman’s correlation coefficient between NIH funding and accepted ASC abstracts between 2016 and 2023.

| Year | Spearman's Correlation Coefficient | p-value |
| --- | --- | --- |
| 2016 | 0.451 | $p < 0.001$ |
| 2017 | 0.533 | $p < 0.001$ |
| 2018 | 0.494 | $p < 0.001$ |
| 2019 | 0.443 | $p < 0.001$ |
| 2020 | 0.518 | $p < 0.001$ |
| 2021 | 0.474 | $p < 0.001$ |
| 2022 | 0.521 | $p < 0.001$ |
| 2023 | 0.506 | $p < 0.001$ |

**Supplementary Table 2:**
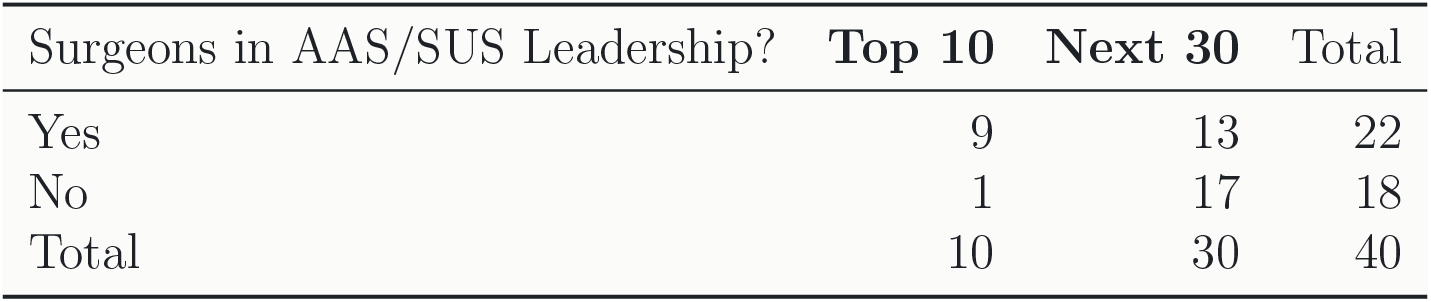
Odds ratio measuring the association between high abstract acceptance and leadership representation in AAS or SUS among top 40 institutions by 2024 ASC abstract acceptances.

